# Atypical Polycystic Kidney Disease in Individuals Heterozygous for *ALG8* Protein-truncating variants

**DOI:** 10.1101/2022.07.13.22277451

**Authors:** Benjamin Apple, Gino Sartori, Bryn Moore, Kiran Chintam, Gurmukteshwar Singh, Prince Mohan Anand, Natasha Strande, Tooraj Mirshahi, William Triffo, Alex Chang

## Abstract

**Background:** Heterozygous *ALG8* variants have previously been associated with polycystic liver disease (PLD) with or without kidney cysts. A clear-cut relationship between application of PKD diagnostic criteria and kidney manifestations of *ALG8* variants remains to be described. We therefore sought to determine whether *ALG8* protein-truncating variant (PTV) heterozygotes are at increased risk of polycystic kidney disease (PKD).

**Methods:** We identified participants heterozygous for pathogenic (P) and likely pathogenic (LP) *ALG8* (NM_024079.5) PTVs described in ClinVar from the Geisinger-Regeneron DiscovEHR MyCode study, an unselected health system-based cohort linked to electronic health records. *ALG8* PTV heterozygotes were matched 1:1 to non-heterozygote family members by age at time of imaging (within 10 years) and sex. Phenotypes were assessed by International Classification of Disease (ICD) codes, chart review, and imaging, which was reviewed by a blinded radiologist. Imaging diagnosis of PKD was defined as ≥4 kidney cysts on an abdominal ultrasound or computed tomography. Secondary outcomes included bilateral renal cysts, and ≥1 liver cyst.

**Results:** Out of 174,418 participants in MyCode,103 participants (mean age 56.7 years) were heterozygous for an *ALG8* P/LP variant: p.Arg364Ter (n=86), p.Arg41Ter (n=7), p.Arg179Ter (n=9), and c.368+2T>G (n=2). None of the *ALG8* P/LP variant heterozygotes had an ICD diagnosis of PKD or PLD. Out of 51 participants ≥40 years of age with available imaging, 51% had ≥4 renal cysts and 14% had ≥1 liver cyst. After matching 23 *ALG8* P/LP variant heterozygotes with 23 related non-heterozygotes by age and imaging modality, *ALG8* P/LP heterozygotes had higher prevalence of 4+ kidney cysts (48% versus 9% in non-heterozygotes; p=0.007) and bilateral kidney cysts (61% vs. 17%; p=0.006).

**Conclusions:** Our study demonstrates that patients heterozygous for *ALG8* P/LP variants are at increased risk of PKD on imaging but not by ICD diagnosis codes. Additional studies are needed to determine whether *ALG8* P/LP heterozygotes are at increased risk of kidney failure.

## Introduction

While *PKD1* and *PKD2* variants account for the majority of autosomal dominant polycystic kidney disease (ADPKD), several other genes have been implicated, including *GANAB, ALG9*, and *DNAJB11*.^1-6^ These genes are involved in the endoplasmic-reticulum processing and maturation of the *PKD1* product polycystin-1.^1,2^ Polycystin-1 is expressed on the renal tubular epithelium and hepatic bile duct epithelium and abnormal polycystin-1 expression leads to polycystic kidney and liver disease.^4,7,8^ Regarding ADPKD, non-*PKD1*/2 variant heterozygotes present with heterogenous and less severe presentations of polycystic kidney disease compared to the traditional *PKD-1/2* ADPKD presentation of enlarged polycystic kidneys and early-onset end-stage kidney disease (ESKD).^1,3,9,10^ The rapidly growing field of nephrogenetics continues to expand on the clinical presentations and associated kidney outcomes of atypical ADPKD.

The *ALG8* (asparagine-linked glycosylation 8) gene, located on chromosome 11q14.1, encodes an endoplasmic reticulum resident protein responsible for glycosylation of proteins such as polycystin-1.^2,11^ *In vitro* experiments show that *ALG8* knockout cells express reduced polycystin-1 glycosylation and total polycystin-1 levels, which are returned to normal levels upon re-expressing the *ALG8* gene.^2^ Decreased levels of functional polycystin-1 within the kidney tubular and liver bile duct epithelium may explain the potential associations of *ALG8* variants with ADPKD and Autosomal Dominant Polycystic Liver Disease (PCLD), respectively.^2,9^ A number of clinical case reports describe homozygous and compound heterozygous *ALG8* variant presentations in the congenital disorder of glycosylation, CDG-Ih.^12-14^ CDG-Ih is a multi-organ disease that can include edema, protein-losing enteropathy, seizures, ataxia, coagulopathy, elevated transaminases, cataracts, and renal tubular dysgenesis, resulting in neonatal or infantile death.^12,14^

In a cohort of 159 unrelated individuals with PCLD, Besse et al. identified 5 individuals with *ALG8* variants including p.Arg364Ter (n=3), p.Arg179Ter (n=1), and a tenth exon splice variant (n=1).^2^ These patients had ≥10 liver cysts, and four of five individuals presented with kidney cysts, ranging from one to nine cysts.^2^ In another cohort of 212 patients with ADPKD who underwent a gene panel analysis that included *PKD1, PKD2*, and 14 other cystogenes, there was 1 proband with a heterozygous *ALG8* p.Leu149Arg variant who presented with renal and hepatic cysts with normal kidney function.^15^ The proband’s mother also carried the same variant and had multiple renal and hepatic cysts, and computational predictions suggested the variant was pathogenic.^15^ Although a few ClinVar submissions have identified additional patients with *ALG8*-related liver and kidney cysts, no other peer-reviewed studies have investigated the cystic kidney or liver phenotype spectrum of monoallelic *ALG8* variant presentations.

As prior studies examined selected cohorts of participants with either PCLD or ADPKD, knowledge of the full phenotypic spectrum of heterozygous *ALG8* variants remains unknown. Given the heterogeneity in the genetic causes of PKD, next-generation sequencing (NGS) is becoming an increasingly used tool for the diagnosis of genetic kidney disease and specifically PKD; several panels include *ALG8*.^1,5^ However, the ClinGen Kidney Cystic and Ciliopathy Disorders Gene Curation Expert Panel (Clinical Genome Resource. https://search.clinicalgenome.org/kb/affiliate/10066?page=1&size=25&search= [12/17/21].) currently has classified the level of evidence for *ALG8* in causing polycystic liver disease with or without kidney cysts as limited.^16^

In this study we examine the phenotypic spectrum of pathogenic/likely pathogenic *ALG8* protein truncating variants (PTVs) using data from the Geisinger-Regeneron DiscovEHR study. Kidney phenotypes of *ALG8* PTV heterozygotes and related non-heterozygotes were characterized by ICD code diagnosis, chart review, and imaging review. We hypothesized that rare *ALG8* PTVs would be associated with increased risk of cystic kidney disease.

## Methods

### Study Population

The Geisinger Institutional Review Board approved this study. Informed consent was waived as participants were previously consented in the MyCode− Community Health Initiative participants as part of the Geisinger-Regeneron DiscovEHR collaboration.^17^ For this study we included adults in MyCode who had whole exome sequencing, and a variant that was classified as pathogenic or likely pathogenic previously in Clinvar. We required genotype quality >20 and alternative allele >3.

We then identified genetically-related family members (1^st^, 2^nd^, or 3^rd^ degree) of *ALG8* PTV heterozygotes, using PRIMUS.^18^ These non-heterozygote relatives of *ALG8* PTV heterozygotes served as a control group.

### Whole exome sequencing

As previously described, WES was performed in collaboration with Regeneron Genetics Center.^17^ Probes from NimbleGen (VCRome) or a modified version of the xGEN probe from Integrated DNA Technologies (IDT) were used for target sequence capture.^19^ Sequencing was performed by paired end 75bp reads on either an Illumina HiSeq2500 or NovaSeq. Coverage depth was sufficient to provide more than 20% coverage over 85% of the targeted bases in 96% of the VCR samples and 90% coverage for 99% of IDT samples. Alignments and variant calling were based on GRCh38 human genome reference sequence.

We included variants that were listed in Clinvar at least once as pathogenic or likely pathogenic (any number of Clinvar stars) and were start-loss, frameshift or early termination/stop-gain of the encoded protein.

### Phenotyping

Phenotyping was performed following similar procedures as done in a prior study that confirmed *ALG9* as a cause of cystic liver and kidney disease.^9^ We used data from the electronic health record (EHR) to ascertain whether participants had International Classification Diseases (ICD) diagnosis codes for polycystic kidney disease (Q61.2, Q61.3, 753.13, 753.12), acquired kidney cysts (N28.1, 593.2), cystic kidney disease (Q61.9, 753.10), liver cystic disease (Q44.6, 573.8, 751.61). cerebral aneurysm (437.3, I67.1), nephrolithiasis (592.*, 594.*, 274.11, N20), dialysis and kidney transplant (see supplement for full set of ICD codes).

To enhance genotype-phenotype analyses for *ALG8*, additional chart review was performed on all participants by at least 2 reviewers, including a nephrologist. Chart review was done by at least 1 nephrologist with focus on kidney imaging data, cerebral aneurysms, history of dialysis, transplant, and family history of ADPKD or cerebral aneurysms. An independent, blinded radiologist (GS) reviewed imaging data for the entire cohort with difficult calls reviewed with a senior radiologist (WT).

### Outcomes

The primary outcome was having ≥ 4 cysts on abdominal ultrasound, computed tomography (CT), or magnetic resonance imaging (MRI) similar to our prior work.^9^ We used a ≥ 4 cyst cutoff as a study of health organ donors found that among adults > age 50 years of age 1, 2, or 3 cysts seen in 26%, 9,8%, and 4.3% of healthy organ donors while only 1.2% had ≥ 4 cysts.^20^ Secondary outcomes included: 1) ≥ 4 cysts or too small to characterize (TSTC) hypodense lesions; 2) bilateral kidney cysts; 3) bilateral kidney cysts or TSTCs; 4) nephrolithiasis (clinical history or on imaging); 5) last eGFR; 6) last eGFR < 60 or ESKD.

### Statistical Analyses

First, we examined heterozygotes of *ALG8* P/LP PTVs and compared their characteristics (sociodemographic, ICD coded definitions, imaging availability, eGFR) to related non-heterozygotes. For kidney and liver cystic ICD code diagnoses and eGFR outcomes, we conducted logistic regression and linear regression for dichotomous and continuous outcomes, respectively. Models were adjusted for age and sex with clustering by family.

Our primary analyses focused on a matched subcohort of *ALG8* P/LP PTV heterozygotes and related non-heterozygotes above the age of 30 who had complete imaging of both kidneys by either CT or MRI. Heterozygotes were matched 1:1 on age quartile and imaging modality (CT with IV contrast, CT without IV contrast, or MRI) with participants in the non-heterozygote relative cohort. Fisher t test was used to examine whether *ALG8* heterozygotes were at increased risk for kidney and liver outcomes. In addition, we conducted a sensitivity analysis that included participants of <u>any age</u> with complete imaging of both kidneys by either CT or MRI, matched 1:1 with non-heterozygotes by age quartile and imaging modality. P values <0.05 were considered statistically significant, and all analyses were conducted using Stata/MP 15.1 (College Station, TX).

## Results

### Characteristics of *ALG8* PTV Heterozygotes and Related Non-heterozygotes

Out of 173,585 participants, there were 229 participants with an *ALG8* PTV, including 103 participants from 85 families who had a PTV that was listed as Pathogenic (P) or Likely Pathogenic (LP) in Clinvar (**Figure 1**). Most participants heterozygous for an *ALG8* P/LP PTVs had Arg364Ter (n=85), followed by Arg179Ter (n=9), Arg41Ter (n=7), and one individual had a splice-donor variant c.368+T2>G.

**Figure 1.**
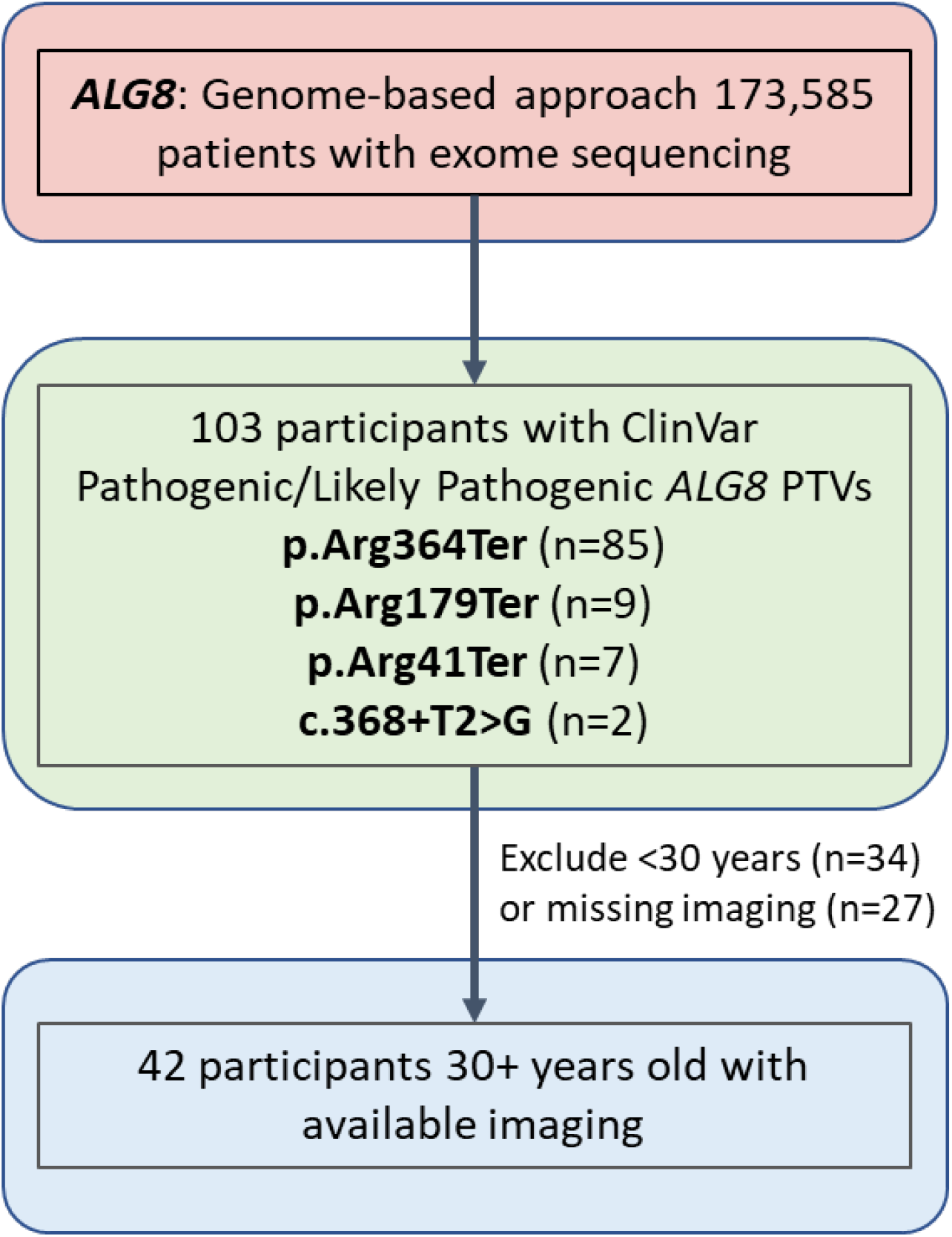
Flowchart.

Within our research cohort, these 103 participants had 55 non-heterozygous relatives. Age (56.5 vs. 58.4 y) and female sex (55.3% vs. 60.0%) were similar between *ALG8* heterozygotes and related non-heterozygotes (**Table 1**). Availability of CT imaging (44.5% and 56.4%) and MRI imaging (3.9% and 10.9%) was similar in *ALG8* heterozygotes and non-heterozygotes.

**Table 1.** Characteristics of *ALG8* P/LP PTV Variant Heterozygotes and related non-Heterozygotes.

|  | **All P/LP Variant heterozygotes (n=103)** | **Related non-heterozygotes (n=55)** |
| --- | --- | --- |
| Age, y | 56.4 (19.9) | 58.4 (17.9) |
| Female, % | 57 (55.3) | 33 (60.0) |
| Self-reported family hx of PKD | 2 (1.9) | 1 (1.8) |
| eGFR available, % | 93 (90.3) | 53 (96.4) |
| Last eGFR, ml/min/1.73m<sup>2</sup> | 80.6 (23.7) | 78.0 (27.5) |
| ESKD, % | 0 | 1 (1.8) |
| Nephrolithiasis, ICD code | 10 (9.7) | 7 (12.7) |
| Nephrolithiasis, ICD code or on imaging | 29 (28.2) | 12 (21.8) |
| ADPKD ICD code | 0 | 0 |
| Cystic kidney disease ICD code | 2 (1.9) | 1 (1.8) |
| Liver cystic disease ICD code | 0 | 0 |
| CT imaging available |  |  |
| No | 46 (44.7) | 23 (41.8) |
| Yes | 46 (44.5) | 31 (56.4) |
| Report only* | 11 (10.7) | 1 (1.8) |
| MRI imaging available, % | 4 (3.9) | 6 (10.9) |
| US - reported left kidney length | N=29<br>11.3 (2.1) | N=23<br>11.5 (1.6) |
| US - reported right kidney length | N=48<br>11.0 (1.7) | N=33<br>11.0 (1.2) |

*ALG8* PTV heterozygotes were not at increased risk of ICD code-diagnosed ADPKD (0% vs. 0%), cystic kidney disease (2% vs. 2%), cystic liver disease (0% vs. 0%), compared to related non-heterozygotes **(Table 1)**. *ALG8* PTV heterozygotes also had no significant differences in prevalence of eGFR <60 ml/min/1.73m^2^ (20.8% vs. 22.6%), ESKD (0% vs. 1.8%), or nephrolithiasis (28.2% vs. 21.8%; p>0.05 for all comparisons in adjusted analyses).

There was no significant difference in right kidney length (11.0 vs. 11.0 cm) or left kidney length (11.3 vs. 11.5 cm) between heterozygotes and non-heterozygotes. Images are shown for Arg364Ter heterozygotes, Arg179Ter heterozygotes, Arg41Ter heterozygotes, and the splice-donor variant (**Figures 1-3)**. The most severe cystic kidney disease presentation was found in a 71-75 year old male carrying a p.Arg41Ter *ALG8* variant. He presented with numerous bilateral renal cysts, with multiple large exophytic cysts and thin septations. There was no clinical history of nephrolithiasis nor ESKD in this patient. Additionally, this *ALG8* PTV heterozygote presented with multiple small hepatic cysts.

**Figure 2.**
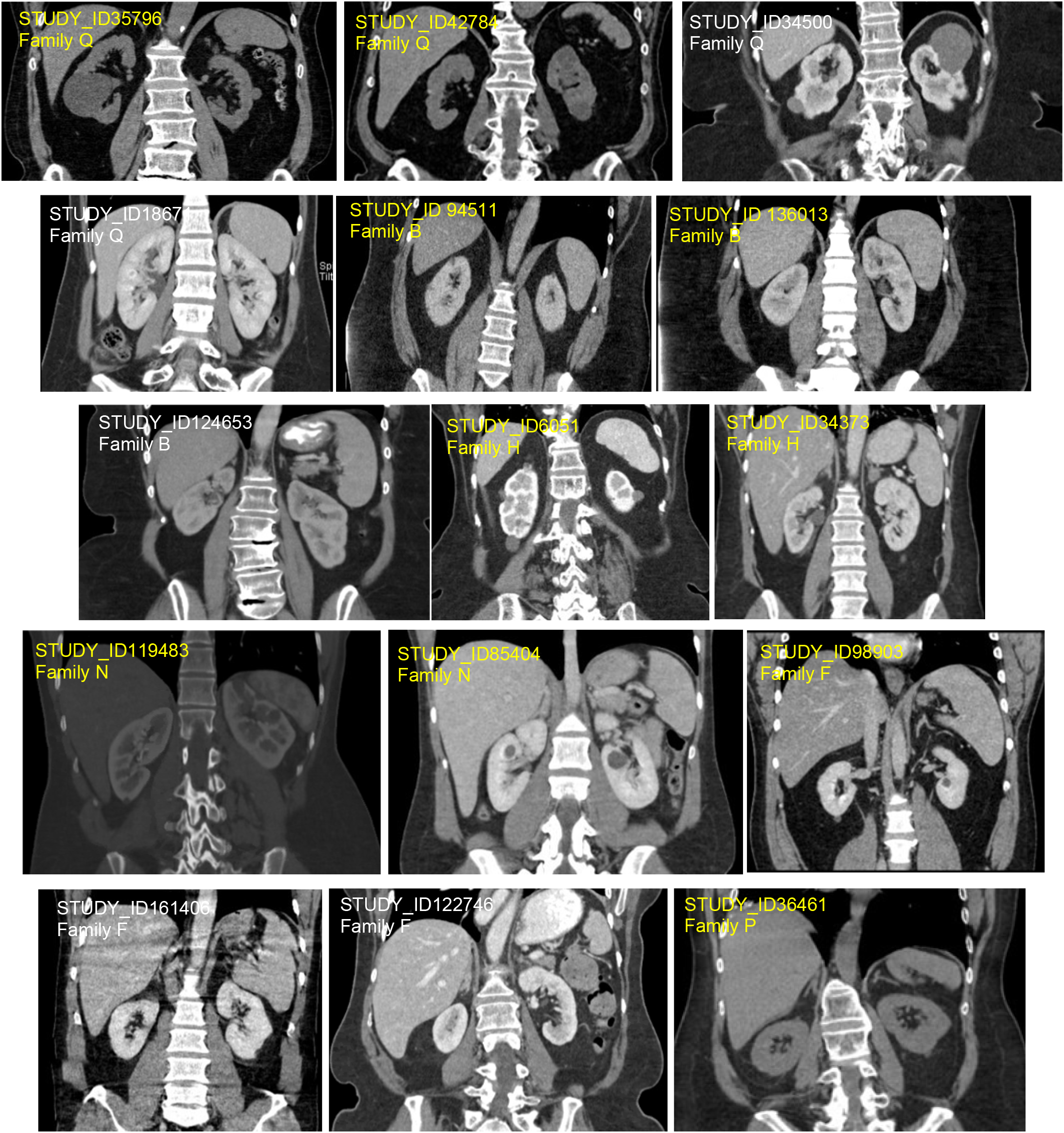
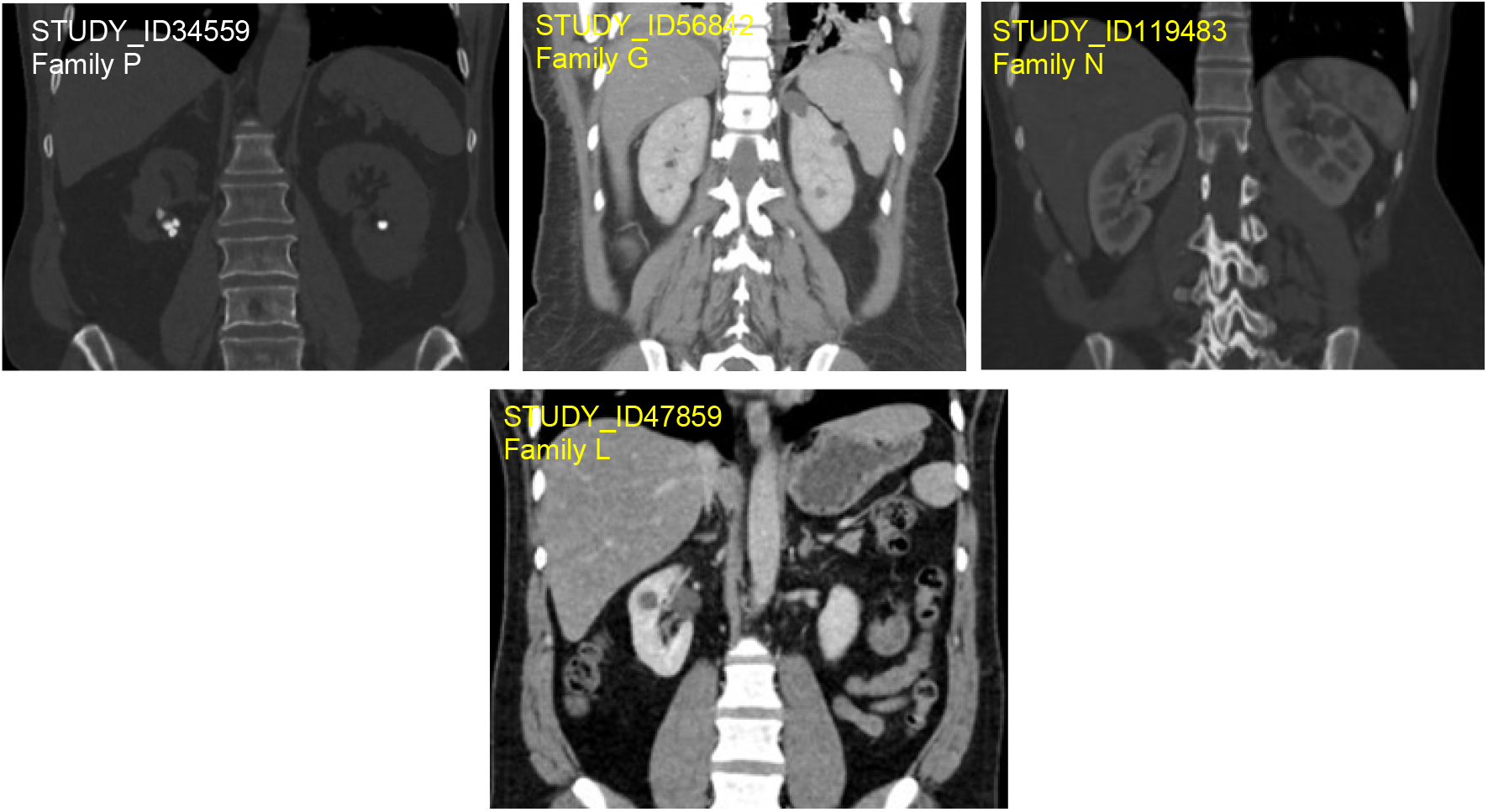
Radiologic Imaging of Families with *ALG8* p.Arg364Ter Heterozygotes. Representative imaging collected from 19 *ALG8* p. Arg364Ter heterozygotes from 9 families.

**Figure 3.**
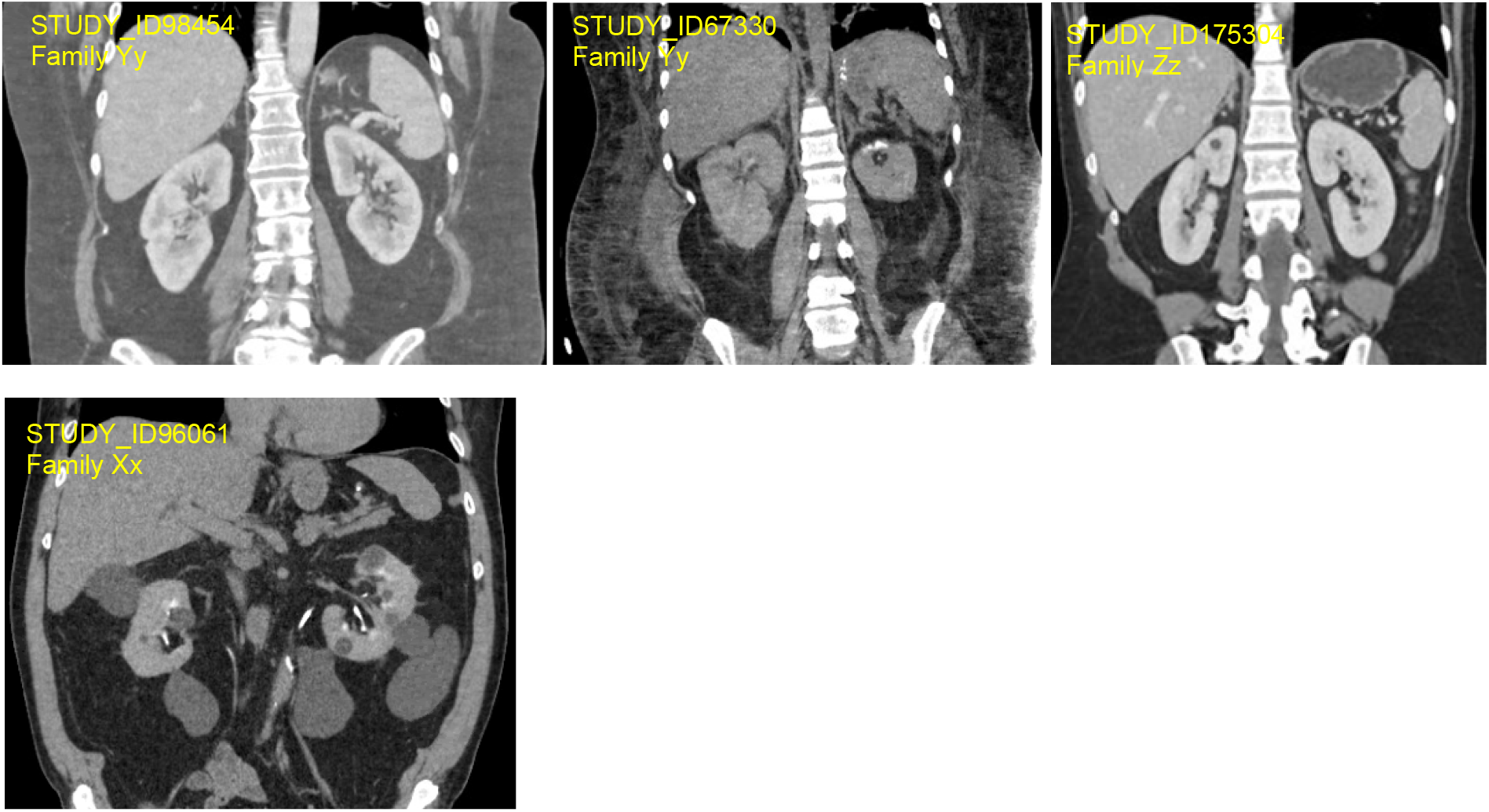
Radiologic Imaging of a Family with *ALG8* p.Arg179Ter Heterozygotes and an individual with ALG8 Arg41Ter. The top row shows representative imaging collected from 3 *ALG8* p.Arg179Ter heterozygotes from 2 families. The bottom row shows imaging from an individual heterozygous for ALG8 Arg41Ter.

### Kidney and Hepatic Cysts among heterozygotes 30 years or older with CT or MRI and matched cohort of related non-heterozygotes

Out of 42 *ALG8* PTV heterozygotes and 28 related non-heterozygotes who were at least 30 years of age and had CT or MRI imaging, a total of 26 heterozygotes were able to be matched 1:1 to 26 non-heterozygotes on age quartile and imaging modality. Mean age at time of imaging was 58.0 and 58.6 years for matched heterozygotes and non-heterozygotes (**Supplemental Table 1**). The most common imaging modality used for evaluation was CT with IV contrast (85%), then CT without IV contrast (12%), and then MRI without contrast (4%). *ALG8* heterozygotes had increased prevalence of ≥4 kidney cysts (57.7% vs. 7.7%), ≥4 kidney cysts or TSTCs (73.1% vs. 19.2%), bilateral renal cysts (69.2% vs. 15.4%), and bilateral renal cysts or TSTCs (84.6% vs. 23.1%; p<0.001 for all comparisons) (**Table 2**). There was no significant difference in prevalence of liver cysts (11.5% vs. 7.7%), liver cysts/TSTCs (23.1% vs. 15.4%), and nephrolithiasis (47.8% vs. 34.8%) between matched *ALG8* heterozygotes and non-heterozygotes.

**Table 2.** Kidney and Liver Cystic Phenotypes of matched *ALG8* P/LP PTV heterozygotes and non-heterozygotes 30+ years of age with CT or MRI imaging.

|  | ***ALG8* P/LP variant heterozygotes (n=26)** | **Non-heterozygotes (n=26)** | **P value** |
| --- | --- | --- | --- |
| ≥ 4 kidney cysts | 15 (57.7%) | 2 (7.7%) | <0.001 |
| ≥ 4 kidney cysts or TSTCs | 19 (73.1%) | 5 (19.2%) | <0.001 |
| Bilateral kidney cysts | 18 (69.2%) | 4 (15.4%) | <0.001 |
| Bilateral kidney cysts or TSTCs | 22 (84.6%) | 6 (23.1%) | <0.001 |
| ≥1 Liver cyst | 3 (11.5%) | 2 (7.7%) | 1.0 |
| ≥1 Liver cyst or TSTCs | 6 (23.1%) | 4 (15.4%) | 0.7 |
| Nephrolithiasis | 14 (53.4%) | 8 (30.1%) | 0.2 |

Heterozygotes were matched 1:1 to individuals in the non-heterozygote relative cohort by age quartile, sex, and imaging modality (CT with IV contrast (n=22 pairs); CT without IV contrast (n=3 pairs), MRI without contrast (n=1 pair).

Findings were similar in a sensitivity analysis that also included individuals in the cohort under the age of 30 with CT or MRI imaging (**Supplemental Table 2**). Prevalence of kidney and liver phenotypes for each of the *ALG8* variants are shown in **Supplemental Table 3**.

### Variability of presentations of *ALG8* Arg364Ter heterozygotes within families

There were 14 families with 2 or more Arg364Ter heterozygotes. Substantial variability in presentation and phenotypic severity was noted among heterozygotes within families. For example, Family B included 3 heterozygotes including a 51–55-year-old with bilateral renal cysts and multiple TSTCs in each kidney and their 31-35-year-old child who had no kidney cysts or TSTCs on IV contrast CT, and a 61-65-year-old cousin with multiple TSTCs in each kidneys (**Figure 2**). Family Q included 2 heterozygotes both with multiple kidney cysts and TSTCs bilaterally **(Figure 2**). Family F included 2 heterozygotes, including a 41–45-year-old with bilateral renal cysts and their 76-80-year-old parent with clinical history of nephrolithiasis but no evidence of kidney cysts on ultrasound (**Figure 2**). Family N included 2 heterozygotes including a 41-45-year-old with bilateral kidney cysts and multiple TSTCs, and their 26–30-year-old child with multiple, bilateral renal cysts (**Figure 2**).

## Discussion

In this study with blinded phenotyping and matched controls, we demonstrate that *ALG8* PTVs increase the risk of kidney cysts with a renal phenotype that appears to be weaker than traditional ADPKD and more akin to the atypical ADPKD associated with *ALG9* and *DNAJB11*.^1,3,5,9^ Despite the high kidney cyst phenotype prevalence among *ALG8* PTV heterozygotes, our study did not find increased frequencies of CKD or ESKD among variant heterozygotes compared to non-heterozygotes. Additional research with larger cohorts of *ALG8* PTV heterozygotes are needed to determine whether these individuals with atypical PKD are at risk for subsequent kidney function decline and ESKD.

Surprisingly, our study did not show a significant association between *ALG8* PTV heterozygotes and polycystic liver phenotypes, as was shown in prior studies of *ALG8* variant heterozygotes.^2^ The association between *ALG8* variants and PLCD has been previously determined by studies that focused on unresolved ADPKD or PLD cases.^2^ As such, prior studies provide a glimpse of the full phenotypic spectrum of disease. By examining a largely unselected health system-based cohort using a genotype-first approach, we are able to provide insights on penetrance, phenotypic severity, and the full spectrum of clinical presentations associated with *ALG8* gene variants.

Our study adds evidence that alterations in enzymes involved in endoplasmic reticulum processing of polycystin-1 are linked to polycystic kidney disease phenotypes, albeit less severe than truncation mutations in *PKD1*.^1,3,6,9^ Given the heterogeneous severity of polycystic kidney disease in our cohort of *ALG8* PTV heterozygotes, the phenotypic severity may depend on a number of additional genetic and environmental factors. Even within families, the cystic kidney disease phenotypes varied widely among relatives carrying *ALG8* PTVs and was often unrecognized in terms of self-described family history in the EHR and lack of ICD diagnosis codes for ADPKD. In contrast of the phenotypic variability and lack of PKD family history seen within *ALG8* PTV families, individuals carrying *PKD1* and *PKD2* gene variants typically all develop a large quantity of bilateral renal cysts with multiple family members progressing to ESKD.^1,5^ Further investigation and identification of other genes associated with polycystic kidney disease will give a more complete understanding of contributions of various gene variants to the development of polycystic kidney disease.

The identification of gene variants may yield potential therapeutic targets for cystic kidney disease by specific genotype. No curative drug therapies currently exist for PKD, although the V2 receptor antagonist, Tolvaptan, has been approved by the FDA to slow renal decline in rapidly progressing ADPKD.^21,22^ Advances in the understanding of the genetic basis of PKD have led to the investigation of various upstream targets of cystogenesis, including vasopressin-2 receptor (AVPR2), EGFR/ErB2, Beta-1 integrin receptor, and c-SRC.^23-25^ It is unknown whether heterozygotes of *ALG8* PTVs would benefit from tolvaptan although tolvaptan is typically indicated only for patients with elevated total kidney volume using the Mayo imaging classification (Mayo Class 1C, 1D, 1E).^22,26,27^ While we did not have uniform imaging to calculate Mayo imaging classification on patients, we found that kidneys were not enlarged for the vast majority of the *ALG8* PTV heterozygotes so most in our study would not qualify from tolvaptan.

A great strength of our study is that we used an unselected patient population to gain a better sense of penetrance, phenotypic severity, and phenotypic spectrum of polycystic disease in *ALG8* PTV heterozygotes. Studies repeatedly show that phenotypic severity is lower in unselected cohorts than in studies examining populations of patients presenting clinically with the disease.^28-31^ In addition, we use related family members of *ALG8* PTV heterozygotes as the comparison group, which reduces residual confounding since family members will share much of the same environmental and background genetic risk factors.

A limitation of the study is that we used routinely collected EHR data rather than rigorously collected research data. Regardless, the proportion of participants with imaging was similar in both groups, and our analysis strategy may be conservative since non-heterozygote participants with kidney imaging are expected to be more likely to have a kidney-related problem compared to those who have no imaging. Use of EHR data could also be considered a strength as this allowed us to examine the full phenotypic spectrum using a genotype-first approach. By limiting our study population to a single healthcare system, the population demographics was limited to a mostly white population, reflecting characteristics of the overall Geisinger population. Follow-up research in other large exome sequencing studies with more diverse populations is needed.

As abdominal imaging has become quite routine in the care of patients presenting with abdominal complaints and for many other reasons, there are many incidental findings of kidney cysts, which could prompt additional anxiety for ADPKD and risk of ESKD. Our study provides important data showing that *ALG8* PTV heterozygotes may have increased risk for mild cystic kidney disease with far fewer consequences compared to patients with ADPKD who have *PKD1* or *PKD2*.^1,5^ There is still much more data to be gathered on the relationship between *ALG8* PTVs and kidney outcomes to confidently apply these findings to *ALG8* PTV heterozygotes with kidney cysts. Additional research is needed to understand what additional risk factors may cause certain individuals with *ALG8* PTVs to develop more severe ADPKD and advanced CKD.

In conclusion, our study suggests that *ALG8* PTVs are associated with increased risk of kidney cysts but not cystic liver disease or ESKD. This adds to the expanding list of genes associated with polycystic kidney and liver diseases.

## Data Availability

Deidentified data produced in the present study are available upon reasonable request to the authors.

