## Supplemental Table 1 for "Atypical Polycystic Kidney Disease in Individuals Heterozygous for *ALG8* Protein-truncating variants"

**Supplemental Table 1. Characteristics of *ALG8* P/LP PTV heterozygotes and matched related non-heterozygotes 30+ years of age with CT or MRI imaging**

| | Heterozygotes (n=26) | Non-heterozygotes (n=26) | P value |
|---|---|---|---|
| Age at imaging, y | 58.0 (16.0) | 58.6 (16.2) | 0.9 |
| Female, % | 15 (57.7) | 13 (50.0) | 0.8 |
| eGFR available, %<br>Last eGFR, ml/min/1.73m$^2$ | 23 (88.5)<br>76.3 (24.0) | 26 (100)<br>73.8 (24.9) | 0.7 |
| ESKD, % | 0 | 0 | - |
| Nephrolithiasis, ICD code | 8 (72.7) | 20 (48.8) | 0.2 |
| Nephrolithiasis, ICD code or on imaging | 14 (60.9) | 14 (48.3) | 0.4 |
| Self-reported family hx of PKD | 1 (3.9) | 1 (3.9) | 1.0 |
| ADPKD ICD code | 0 | 0 | - |
| Cystic kidney disease ICD code | 1 (3.9) | 1 (3.9) | 1.0 |
| Acquired kidney cyst | 2 (7.7) | 1 (3.9) | 1.0 |
| Liver cystic disease ICD code | 0 | 0 | - |
| Imaging modality<br>CT with IV contrast<br>CT without IV contrast<br>MRI without IV contrast | <br>22 (84.6)<br>3 (11.5)<br>1 (3.9) | <br>22 (84.6)<br>3 (11.5)<br>1 (3.9) | 1.0 |
