## Supplemental Table 2 for "Atypical Polycystic Kidney Disease in Individuals Heterozygous for *ALG8* Protein-truncating variants"

**Supplemental Table 2. Kidney and Liver Cystic Phenotypes of *ALG8* P/LP PTV heterozygotes and matched related non-heterozygotes with CT or MRI imaging**

| | **All P/LP Variant heterozygotes (n=36)** | **Related non-heterozygotes (n=24)** | **P value** |
|---|---|---|---|
| ≥ 4 kidney cysts | 12 (41.4) | 2 (6.9) | 0.005 |
| ≥ 4 kidney cysts or TSTCs | 19 (65.5) | 5 (17.2) | <0.001 |
| Bilateral kidney cysts | 17 (58.6) | 4 (13.8) | 0.001 |
| Bilateral kidney cysts or TSTCs | 21 (72.4) | 7 (24.1) | 0.001 |
| Liver cysts | 2 (6.9) | 2 (6.9) | 1.0 |
| Liver cysts or TSTCs | 8 (27.6) | 4 (13.8) | 0.3 |
| Nephrolithiasis | 14 (48.3) | 9 (31.0) | 0.3 |

Heterozygotes were matched 1:1 to individuals in the non-heterozygote relative cohort by age sextile, sex, and imaging modality (CT with IV contrast (n=25 pairs); CT without IV contrast (n=4 pairs)
