## Supplemental Table 3 for "Atypical Polycystic Kidney Disease in Individuals Heterozygous for *ALG8* Protein-truncating variants"

**Supplemental Table 3. Kidney and Liver Cystic Phenotypes of *ALG8* P/LP PTV heterozygotes with CT or MRI imaging, by Variant**

| | **p.Arg364Ter<br>Age ≥30: n=34<br>Age <30: n=4** | **p.Arg179Ter<br>Age ≥30: n=2<br>Age <30: n=1** | **p.Arg41Ter<br>Age ≥30: n=5<br>Age <30: n=0** | **c.368+T2>G<br>Age ≥30: 1<br>Age <30: 0** |
|---|---|---|---|---|
| Consequence | Stop-gained | Stop-gained | Stop-gained | Splice donor |
| Clinvar classification | Pathogenic<br>8 submission<br>2 stars | Pathogenic<br>1 submission<br>0 stars | Pathogenic<br>3 submissions<br>2 stars | Likely pathogenic<br>1 submission<br>1 star |
| Conditions listed in Clinvar | Polycystic liver disease 3 with kidney cysts (AD); Congenital disorder of glycosylation type 1H (AR) | Polycystic liver disease 3 with kidney cysts (AD) | Congenital disorder of glycosylation type 1H (AR) | Congenital disorder of glycosylation type 1H (AR) |
| ≥ 4 kidney cysts | Age ≥30: 18/34 (52.9%)<br>Age <30: 1/4 (25%) | Age ≥30: 0/2<br>Age <30: 0/1 | Age ≥30: 2/5 (60%) | Age ≥30: 1/1 (100%) |
| ≥ 4 kidney cysts or TSTCs | Age ≥30: 25/34 (73.5%)<br>Age <30: 2/4 (50%) | Age ≥30: 2/2 (100%)<br>Age <30: 0/1 ( | Age ≥30: 3/5 (60%) | Age ≥30: 1/1 (100%) |
| Bilateral kidney cysts | Age ≥30: 23/34 (67.7%)<br>Age <30: 2/4 (50%) | Age ≥30: 1/2 (50%)<br>Age <30: 0/1 | Age ≥30: 3/5 (60%) | Age ≥30: 1/1 (100%) |
| Bilateral kidney cysts or TSTCs | Age ≥30: 27/34 (79.4%)<br>Age <30: 2/4 (50%) | Age ≥30: 2/2 (100%)<br>Age <30: 1/1 (100%) | Age ≥30: 4/5 (80%) | Age ≥30: 1/1 (100%) |
| Liver cysts | Age ≥30: 3/34 (8.8%)<br>Age <30:<br>0/4 | Age ≥30: 0/2<br>Age <30: 0/1 | Age ≥30: 1/5 (20%) | Age ≥30: 1/1 (100%) |

| | | | | |
|---|---|---|---|---|
| Liver cysts or TSTCs | Age ≥30: 10/34 (29.4%)<br>Age <30: 1/4 (25%) | Age ≥30: 0/2<br>Age <30: 0/1 | Age ≥30: ⅕ (20%) | Age ≥30: 1/1 (100%) |
| Nephrolithiasis | Age ≥30: 18/34 (52.9%)<br>Age <30: 1/4 (25%) | Age ≥30: 0/2<br>Age <30: 0/1 | Age ≥30: 3/5 (60%) | Age ≥30: 1/1 (100%) |

This table only includes families where a proband had imaging (complete US, CT, or MRI) above the age of 20.
